# Microbial etiology, antibiotic susceptibility profiles, and multidrug resistance of urinary tract infections at a secondary healthcare facility in Ghana

**DOI:** 10.64898/2026.06.11.26355450

**Authors:** John Kofi Agyapong, Godfred Damalie, Rebecca Dombawel, Noah Akwasi, Yaa Balo, Anabel Acheampong, Prince-Charles Kudzordzi, Prince Nyarko, Dennis Kyei Ofori, Kenneth Bentum Otabil

## Abstract

**Background:** Rising antibiotic resistance challenges empirical therapies for urinary tract infections (UTIs). This study evaluated the microbial etiology, susceptibility profiles, and multidrug resistance (MDR) patterns of uropathogens among outpatients at the Berekum Holy Family Hospital, Ghana.

**Methods:** This cross-sectional study (February–August 2021) screened 263 symptomatic outpatients. Mid-stream urine samples underwent quantitative culture, biochemical identification, and antimicrobial susceptibility testing via the Kirby-Bauer disc diffusion method following the 2021 CLSI guidelines.

**Results:** Significant bacteriuria prevalence was 22.8% (60/263). UTIs predominated in females (78.3%, 47/60; p = 0.1501) and individuals ≥45 years (33.3%, 20/60). Gram-negative rods accounted for 90.0% of isolates, primarily *Escherichia coli* (26.7%), *Citrobacter* spp. (25.0%), and *Enterobacter* spp. (21.7%); *Staphylococcus aureus* (10.0%) was the only Gram-positive pathogen. Extreme phenotypic resistance was observed against piperacillin/tazobactam (98.3%), cefotaxime (93.3%), tetracycline (88.3%), and cefoperazone (85.0%). Conversely, highest therapeutic susceptibilities were retained by amikacin (78.3%), levofloxacin (61.7%), and gentamicin (58.3%).

**Conclusion:** The high prevalence of MDR uropathogens against advanced beta-lactamase inhibitor combinations and cephalosporins necessitates an immediate re-evaluation of regional empirical protocols. Amikacin, levofloxacin, and gentamicin remain viable options prior to culture confirmation. These findings establish a crucial phenotypic baseline to guide localized prescribing policies and regional antimicrobial resistance tracking strategies.

## Background

Urinary tract infections (UTIs) represent a substantial burden within clinical and outpatient settings globally, with approximately 3 million individuals visiting health services for UTIs each year [1]. The clinical manifestations range from uncomplicated localized cystitis to severe, complicated pyelonephritis or systemic urosepsis [2]. UTIs cause substantial morbidity and direct economic strains on healthcare infrastructure, coupled with indirect productivity losses aggregate to billions of dollars annually worldwide [1]. Although viral and fungal etiologies of UTIs are documented, bacterial uropathogens remain the primary drivers of clinical infection [3]. The demographics of the patient including age, gender, hospitalization history, and prior exposure to antimicrobials dynamically influence the epidemiological patterns [3]. However, comprehensive and current epidemiological data remain scarce [1].

The gold standard for accurate UTI diagnosis is microbiological isolation and quantitative determination of specific pathogens in clean-catch mid-stream urine samples, paired with corresponding clinical symptoms [4]. Epidemiological surveys consistently document that community-acquired infections are primarily driven by enteric Gram-negative rods, notably *Escherichia coli, Klebsiella pneumoniae*, and *Citrobacter* species, alongside Gram-positive opportunistic agents like *Staphylococcus saprophyticus* or *Staphylococcus aureus* [5, 6]. Conversely, hospital acquired UTIs display increased etiological complexity, frequently involving problematic pathogens such as *Pseudomonas aeruginosa* and *Proteus mirabilis* [7, 8]. These pathogens display a propensity for developing accelerated resistance secondary to intense institutional selective pressures.

The rapid emergence and dissemination of antimicrobial resistance (AMR) among uropathogens is considered as a global health emergency, as highlighted by surveillance initiatives from the World Health Organization [9]. In low- and middle-income countries (LMICs), including Ghana, this crisis is compounded by infrastructural limitations, a lack of institutionalized diagnostic stewardship, and unregulated access to over-the-counter antimicrobials without formal prescriptions [10, 11]. The resulting expansion of the regional resistome underscores an urgent need for continuous monitoring at local sentinel sites. Indeed, this priority has recently been emphasized within the literature as vital for mapping precise epidemiological baselines within highly vulnerable regional cohorts [12].

In a previous study, we reported on the baseline susceptibility patterns in cutaneous and wound infections in the middle belt of Ghana [13]. However, a critical gap remains with regards to the specific structural selective pressures shaping uropathogen communities [13]. This is necessary because pathogens in the urinary tract operate under distinct diagnostic, anatomical, and empirical treatment regimens compared to those in surface wounds. Thus, localized surveillance and tracking of their unique multi-drug cross-resistance pathways is an absolute public health mandate to prevent clinical treatment failure in UTIs. To mitigate the expansion of localized AMR and preserve therapeutic efficacy, continuous facility-specific surveillance is vital to establish evidence-based guidelines for empirical prescription.

This study evaluated the microbial etiology, antibiotic susceptibility profiles, and multidrug resistance patterns of uropathogens among symptomatic outpatients attending the Berekum Holy Family Hospital in the Bono Region of Ghana. It establishes vital baseline phenotypic multi-drug resistance (MDR) patterns and provide essential regional data to inform global antimicrobial resistance tracking networks while simultaneously optimizing localized empirical prescribing guidelines.

## Methods

### Study design and setting

A facility-based cross-sectional study was implemented from February 21, 2021, to August 28, 2021, at the Berekum Holy Family Hospital. This institution is a secondary healthcare facility that serves as the main hospital in the Berekum Municipality. It is positioned within the North-western sector of the Bono Region, Ghana (latitude 7°20’50’’ N, and longitude 2°20’30’’ W). The facility manages an average annual outpatient department (OPD) volume exceeding 100,000 cases, maintains a 162-bed capacity alongside an active 100-bed maternity unit. It delivers specialized interventions in general surgery, internal medicine, pediatrics, and urology, making it an ideal sentinel site for AMR monitoring.

### Sample size determination and sampling technique

The target sample size was calculated utilizing standard single proportion estimation based on a previously reported baseline UTI prevalence of 22.5% from the Eastern Regional Hospital in Koforidua, Ghana [14]. Employing a 95% confidence interval and a 5% margin of error, the minimum required sample size was determined. A total of 263 outpatients suspected of having a UTI based on initial clinical presentation were consecutively enrolled via a random sampling design during their primary consultation.

### Inclusion and exclusion criteria

Inclusion criteria comprised all ambulatory outpatients presenting with clinical features indicative of a UTI. These included acute dysuria, severe urinary frequency, lower abdominal urgency, or visible hematuria, as well as cases of asymptomatic bacteriuria identified by the attending clinicians. Exclusion criteria strictly applied to pediatric patients below 1 year of age, pregnant women in active labor, and postpartum women within 24 hours of delivery, to exclude confounding physiologic or healthcare-associated variants.

### Sample collection and processing

Following structured explanations provided in the local dialect (Twi), consenting participants were instructed on aseptic techniques to self-collect a 30 mL clean-catch mid-stream urine sample using sterile, wide-mouthed containers. Each sample was uniquely labelled using specific demographic data (age, gender, clinical status, and current empirical treatment). The samples were processed within 1 hour of collection at the microbiology laboratory.

### Culturing and identification of uropathogens

Macroscopic parameters including color and appearance were documented. Quantitative inoculation was performed by streaking a calibrated platinum loopful (0.001 mL) of uncentrifuged urine onto standard Cysteine Lactose Electrolyte-Deficient (CLED) agar plates [15]. Inoculated media were incubated aerobically at 37°C for 18–24 hours. Post-incubation, colony counting was performed, and significant bacteriuria defined by a primary uropathogen count equal to or exceeding 1×10^5 colony-forming units per milliliter (CFU/mL) [15]. Isolated distinct colonies were subjected to standard Gram staining to differentiate cell morphology.

Gram-positive cocci were further differentiated via catalase testing, and coagulase assays profiles. Gram-negative bacilli were identified to the genus level using biochemical tests, including indole production, citrate utilization, urease activity, Triple Sugar Iron (TSI) agar fermentation, and oxidase tests [15].

### Antimicrobial susceptibility testing (AST)

*In vitro* susceptibility profiling was evaluated via the standard Kirby-Bauer disc diffusion technique, strictly following the interpretive criteria of the 2021 Clinical and Laboratory Standards Institute (CLSI) [16]. Well-isolated colonies from 24-hour pure cultures were suspended in sterile normal saline, with turbidity adjusted to match a 0.5 McFarland standard. Using sterile cotton swabs, the suspension was inoculated uniformly across the surface of Mueller-Hinton agar plates. Commercially manufactured antibiotic discs (Biomark Laboratories, India) were applied, consisting of a 12-drug panel: amikacin (30 µg), cefoperazone (75 µg), ceftriaxone (10 µg), ciprofloxacin (5 µg), piperacillin/tazobactam (10 µg), cefotaxime (30 µg), tetracycline (30 µg), nitrofurantoin (300 µg), gentamicin (10 µg), norfloxacin (10 µg), nalidixic acid (30 µg), and levofloxacin (5 µg). Following aerobic incubation at 37°C for 18–24 hours, zone inhibition diameters were measured to the nearest millimeter and stratified as sensitive, intermediate, or resistant.

Multidrug resistance (MDR) was defined as phenotypic non-susceptibility to at least one agent in three or more antimicrobial categories, in accordance with the international consensus criteria established by Magiorakos et al [17]. For the calculation of MDR fractions, non-susceptibility profiles were strictly assessed based on acquired resistance patterns, ensuring that species-specific intrinsic resistance properties did not affect the cumulative multi-class resistance metrics.

### Statistical analysis

Field sheets were reviewed for data integrity, digitized within Microsoft Excel, and subsequently imported into GraphPad Prism version 8.0 for statistical analysis. Categorical demographic metrics and prevalence fractions were expressed as percentages. Cross-tabulations between gender distribution and culture positivity were analyzed via the Fisher’s exact test, with statistical significance declared at a p-value < 0.05. Continuous parameters like age were stated as ranges and median values.

### Ethical consideration and data protection

Formal institutional ethical clearance was granted by the Committee for Human Research and Ethics (CHRE) at the Department of Basic and Applied Biology, School of Sciences, University of Energy and Natural Resources (Reference: CHRE/AP/03/021). Written informed consent (signed or thumbprinted) was taken from all participants prior to sample processing. Confidentially was strictly maintained in accordance with the Ghana Data Protection Act (Act 843, 2012).

## Results

### Socio-demographic characteristics of study participants

Majority of the study participants, 70.7% (186/263) were females (Table 1). Stratification by age groups revealed that young adults aged 18–29 years represented the largest category (29.3%, 77/263), followed closely by adults aged 30–45 years (26.2%, 69/263), children under 18 years (22.4%, 59/263), and the elderly aged 45 years and above (22.1%, 58/263) (Table 1). The median age of all participants was 29.0 years (Interquartile Range [IQR]: 19.0 - 41.0).

**Table 1.** Distribution of study participants by sex and age category.

| **Variable** | **Percentage, (frequency)** |
| --- | --- |
| **Sex** |  |
| Males | 29.3% (77) |
| Females | 70.7% (186) |
| **Age category** |  |
| Children (<18 years) | 22.4% (59) |
| Young (18-29 years) | 29.3% (77) |
| Adults (30-45 years) | 26.2% (69) |
| Old (≥45 years) | 22.1% (58) |

### Prevalence of UTI by sex and age

Microbiological analysis of the 263 urine specimens confirmed significant bacteriuria (≥1×10^5 CFU/mL) in 22.8% (60/263) of outpatients. Insignificant growth (<1×10^5 CFU/mL) was observed in 8.4% (22/263) of the specimens, while 68.8% (181/263) yielded no microbial growth.

Sex-based stratification of confirmed UTI cases showed a substantial concentration among female outpatients, who accounted for 78.3% (47/60) of the infections, compared to 21.7% (13/60) among males (Table 2). However, inferential statistical analysis demonstrated that this variance was not statistically significant within this study population (Fisher exact test; p = 0.1501).

**Table 2.** Prevalence of UTI by age categories.

| Age Group | Males, n+ve (%) | Females, n+ve (%) | Prevalence, % (n/N) [95% CI] |
| --- | --- | --- | --- |
| Children (<18 years) | 3 (5.1%) | 6 (10.2%) | 15.3% (9/59) [8.2 – 26.5%] |
| Young (18-29 years) | 1 (1.3%) | 15 (19.5%) | 20.7% (16/77) [13.2 – 31.1] |
| Adults (30-45 years) | 0 (0.0%) | 15 (21.7%) | 21.7% (15/69) [13.6 -32.8] |
| Old ($\geq 45$ years) | 9 (15.5%) | 11 (19.0%) | 34.5% (20/58) [23.6 – 47.3] |
| **Total** | **13 (4.9%)** | **47 (17.9%)** | **22.8% (60/263) [18.2 – 28.3]** |

When stratified across age categories, the elderly category (≥45 years) demonstrated the highest age-specific infection rate, with 34.5% (20/58) of individuals in this bracket testing positive for significant bacteriuria (Table 2). In terms of the total burden of infection, this elderly category constituted the largest proportion of the confirmed UTI cases at 33.3% (20/60), followed by young adults at 26.7% (16/60), adults at 25.0% (15/60), and children at 15.0% (9/60). The median age of individuals diagnosed with a UTI was 31.0 years (IQR: 24.0 - 58.3).

Notably, gender distribution within these age categories revealed that females consistently predominated across all classes, particularly within the young adult (15/16) and adult (15/15) categories, where they accounted for nearly all cases. On the contrary, in the male category, susceptibility was concentrated among the elderly, with 69.2% (9/13) of all male UTIs occurring in individuals aged 45 years and above (Table 2).

### Distribution of specific bacterial uropathogens

Gram-negative bacilli were the primary drivers of infection, accounting for 90.0% (54/60) of all positive isolates (Table 3). Overall, *E. coli* was the most prevalent uropathogen, accounting for 26.7% (16/60), followed by *Citrobacter* spp. (25.0%, 15/60) and *Enterobacter* spp. (21.7%, 13/60). Other Gram-negative rods identified included *Klebsiella* spp. (6.7%, 4/60), *Providencia* spp. (3.3%, 2/60), *P. aeruginosa* (3.3%, 2/60), and *Shigella* spp. (3.3%, 2/60). *S. aureus* was the sole Gram-positive species isolated, representing 10.0% (6/60) of the positive cultures (Table 3).

**Table 3.**
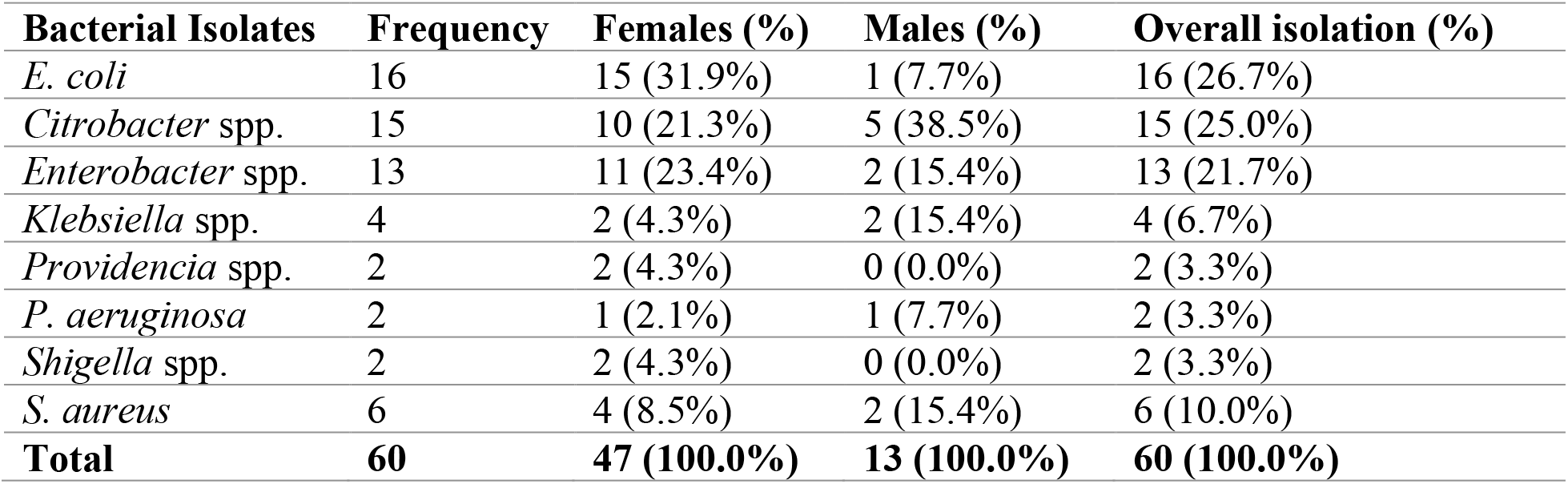
Distribution of specific bacterial uropathogens by sex.

### Antimicrobial susceptibility and resistance profiles

The bacterial isolates displayed extensive *in vitro* resistance to most tested antibiotics (Table 4). Alarming phenotypic resistance was observed against advanced therapeutic combinations, with 98.3% (59/60) of all ambulatory outpatients exhibiting complete clinical non-susceptibility to piperacillin/tazobactam. There was also an overwhelming collapse of third generation cephalosporins including cefotaxime (93.3%) and cefoperazone (85.0%). High resistance rates were also observed for tetracycline (88.3%, 53/60), ceftriaxone (78.3%, 47), and nalidixic acid (71.7%, 43/60). In contrast, amikacin was the most effective agent tested, showing an overall sensitivity rate of 78.3% (47/60) across both Gram-positive and Gram-negative isolates. Other effective choices against Gram-negative bacilli included levofloxacin and gentamicin, which demonstrated sensitivity rates of 61.7% (37/60) and 58.3% (35/60), respectively (Table 4).

**Table 4.** Antimicrobial susceptibility and resistance profiles.

| **Antimicrobial Agent** | **Sensitive, n (%)** | **Resistant, n (%)** |
| --- | --- | --- |
| Piperacillin/Tazobactam (10 µg) | 1 (1.7%) | 59 (98.3%) |
| Cefotaxime (30 µg) | 4 (6.7%) | 56 (93.3%) |
| Tetracycline (30 µg) | 7 (11.7%) | 53 (88.3%) |
| Cefoperazone (75 µg) | 9 (15.0%) | 51 (85.0%) |
| Ceftriaxone (10 µg) | 13 (21.7%) | 47 (78.3%) |
| Nalidixic Acid (30 µg) | 17 (28.3%) | 43 (71.7%) |
| Nitrofurantoin (300 µg) | 21 (35.0%) | 39 (65.0%) |
| Norfloxacin (10 µg) | 26 (43.3%) | 34 (56.7%) |
| Ciprofloxacin (5 µg) | 27 (45.0%) | 33 (55.0%) |
| Gentamicin (10 µg) | 35 (58.3%) | 25 (41.7%) |
| Levofloxacin (5 µg) | 37 (61.7%) | 23 (38.3%) |
| Amikacin (30 µg) | 47 (78.3%) | 13 (21.7%) |

### Prevalence of multi drug resistance (MDR) among uropathogens

We evaluated cumulative cross-resistance profiles across the 6 structural classes tested (including aminoglycosides, penicillin/beta-lactamase inhibitors, cephalosporins, fluoroquinolones, tetracyclines, and nitrofurans) (Table 5). A total of 96.7% (58/60) of the recovered bacterial uropathogens were confirmed to be MDR. Universal MDR saturation (100.0%) was observed across nearly all isolated genera, including *Citrobacter* spp. (n=15), *Enterobacter* spp. (n=13), *Klebsiella* spp. (n=4), *P. aeruginosa* (n=2), *Shigella* spp. (n=2), and *Providencia* spp. (n=2). Notably, 20.0% (12/60) of the total isolates exhibited concurrent resistance to every single drug class evaluated in the study, severely narrowing potential empirical treatment pathways.

**Table 5.**
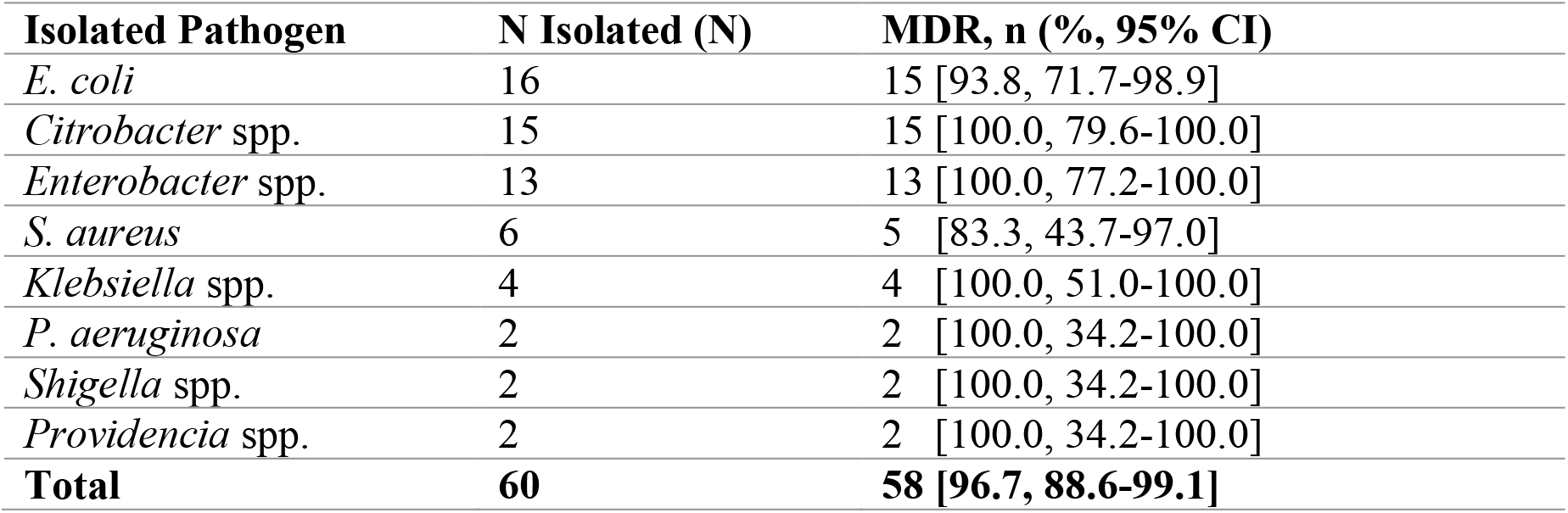
Prevalence of multi drug resistance (MDR) among uropathogens.

| Isolated Pathogen | N Isolated (N) | MDR, n (% , 95% CI) |
| --- | --- | --- |
| *E. coli* | 16 | 15 [93.8, 71.7-98.9] |
| *Citrobacter* spp. | 15 | 15 [100.0, 79.6-100.0] |
| *Enterobacter* spp. | 13 | 13 [100.0, 77.2-100.0] |
| *S. aureus* | 6 | 5 [83.3, 43.7-97.0] |
| *Klebsiella* spp. | 4 | 4 [100.0, 51.0-100.0] |
| *P. aeruginosa* | 2 | 2 [100.0, 34.2-100.0] |
| *Shigella* spp. | 2 | 2 [100.0, 34.2-100.0] |
| *Providencia* spp. | 2 | 2 [100.0, 34.2-100.0] |
| **Total** | **60** | **58 [96.7, 88.6-99.1]** |

## Discussion

This study evaluated the microbial etiology, antibiotic susceptibility profiles, and multidrug resistance patterns of uropathogens among symptomatic outpatients attending the Berekum Holy Family Hospital in the Bono Region of Ghana.

The study revealed a 22.8% prevalence of significant bacteriuria among symptomatic ambulatory outpatients at the Berekum Holy Family Hospital. This prevalence rate aligns closely with the 22.5% reported by Fofana in Koforidua in Ghana [14] and the 21.2% reported by Tadesse et al [18] in Nigeria. However, it is higher than the prevalence reported by Donkor et al [10] in southern Ghana (10.1%) and Labi et al [19] in northern Ghana (18.5%). On the contrary, our findings are lower than those from the Ghana Police Hospital in Accra (31.6%) [5] and the Komfo Anokye Teaching Hospital in Kumasi (34.5%) [20]. These regional differences are likely driven by variations in diagnostic criteria, socioeconomic conditions, and local water and personal hygiene practices [11]. The observed prevalence may also underestimate the true burden, as our conventional culture workflow could not detect atypical fastidious pathogens like *Chlamydia trachomatis, Mycoplasma hominis*, or *Ureaplasma urealyticum*, which require specialized media or molecular techniques [21].

The higher prevalence of UTIs observed in females (78.3%) compared to males (21.7%) is consistent with established epidemiological trends [5, 22, 23]. This disparity is primarily due to anatomical factors, including the shorter female urethra and its proximity to the warm, moist perineal and perianal areas. This facilitates the migration of enteric normal flora into the urinary tract [24, 25]. Cross-tabulating age and sex revealed a high concentration of infections among young females (18–29 years) and older males (≥45 years). In young women, this vulnerability is frequently associated with high sexual activity, the use of spermicides, and mechanical irritation [26]. In older men, the sharp increase in susceptibility (69.2% of all male cases) is typical of aging-related anatomical changes [27]. These include benign prostatic hyperplasia, which leads to urinary stasis and incomplete bladder emptying and favor bacterial colonization [27].

Gram-negative bacilli dominated the etiological profile (90.0%), consistent with prior studies [26, 28]. *E. coli* was the most common isolate (26.7%), followed closely by *Citrobacter* spp. (25.0%) and *Enterobacter* spp. (21.7%). The high prevalence of *Citrobacter* and *Enterobacter* species reflects an expanding reservoir of these opportunistic pathogens within the community. *S. aureus* was the only Gram-positive organism isolated (10.0%). This confirms that while Gram-positive bacteria are less common in uncomplicated UTIs, they remain an important clinical consideration.

The antimicrobial susceptibility profiles revealed worryingly high resistance rates to commonly prescribed empirical antibiotics. The 98.3% resistance rate observed for piperacillin/tazobactam and 93.3% for cefotaxime are particularly concerning. These high rates indicate a widespread distribution of extended-spectrum beta-lactamase (ESBL)-producing strains or other advanced beta-lactamase resistance mechanisms in the community, similar to trends reported across Ghana [29, 5]. Piperacillin/tazobactam is an intravenous antibiotic. Thus, the near-universal resistance among ambulatory outpatients is likely driven by plasmid-mediated co-selection. In co-selection, the genes encoding resistance to a heavily misused, cheap oral drugs could be structurally linked on the same mobile genetic elements or through hyper-expression of chromosomal *AmpC β*-lactamases within the abundant *Citrobacter* and *Enterobacter* isolates [5, 29].

High resistance rates were also observed for tetracycline (88.3%), cefoperazone (85.0%), and ceftriaxone (78.3%). This resistance is likely driven by intense selective pressures resulting from the unregulated over-the-counter availability and frequent use of these affordable oral alternatives [30].

In contrast, amikacin demonstrated exceptional *in vitro* efficacy, with a 78.3% overall sensitivity rate. This high susceptibility aligns with findings in Ghana [31,32] and is likely due to its restricted parenteral administration, which limits community misuse. Levofloxacin (61.7% sensitive) and gentamicin (58.3% sensitive) also showed moderate efficacy, suggesting they remain viable therapeutic options. These findings emphasize the urgent need to establish facility-specific diagnostic stewardship initiatives to replace outdated empirical protocols with evidence-based options.

The observed 96.7% multi-drug resistance (MDR) rate among uropathogens in this setting represents a critical breakdown in local empirical treatment options. This extreme rate of phenotypic cross-resistance is likely driven by the unregulated, over-the-counter availability of broad-spectrum antimicrobials in the community, coupled with a lack of routine local laboratory surveillance [30]. When first-line selections like tetracycline and third generation cephalosporins fail at rates exceeding 75%, clinicians are forced to rely on highly restrictive classes, rapidly exhausting the regional formulary. This further supports our call for re-evaluation of facility specific AMR stewardship programmes to ensure their continuous effectiveness.

We acknowledge the limitation of a lack of molecular confirmation, such as Polymerase Chain Reaction (PCR) assays, to characterize specific resistance genes (e.g., *bla*_CTX-M_, *mecA*, or *vanA*). Consequently, our findings are strictly phenotypic. However, given that phenotypic MDR has reached an absolute saturation point of nearly 100% across key Gram-negative isolates in this hospital cohort, these findings serve as an urgent epidemiological alert. This baseline mapping demonstrates that immediate molecular surveillance is an essential public health priority to identify the plasmids driving this massive regional resistome.

## Conclusion

This study demonstrates a high prevalence of significant bacteriuria (22.8%) among outpatients at the Berekum Holy Family Hospital, driven primarily by Gram-negative enteric bacilli. The isolated uropathogens displayed extensive resistance to commonly used empirical antibiotics, including piperacillin/tazobactam, third generation cephalosporins, and tetracyclines. Amikacin, levofloxacin, and gentamicin emerged as the most effective agents *in vitro*. This study has established a vital baseline phenotypic MDR) pattern that provides essential regional data to inform global antimicrobial resistance tracking strategies while simultaneously optimizing localized empirical prescribing guidelines.

## Data Availability

All data produced in the present study are available upon reasonable request to the authors.

## Declarations

### Ethics approval and consent to participate

The study protocol was formally approved by the Committee for Human Research and Ethics (CHRE) of the erstwhile Department of Basic and Applied Biology, School of Sciences, University of Energy and Natural Resources, Sunyani (Reference: CHRE/AP/03/021). Written informed consent was obtained from all participants or their parents/legal guardians (in the case of minors <18 years), and the participants were free to withdraw from the study at any time without consequence.

### Consent to participate

Written, signed/thumb printed informed consent was sought from all participants ahead of inclusion in the study.

### Declaration of generative AI and AI-assisted technologies in the writing process

During the preparation of this work, the authors used artificial intelligence (AI) and AI-assisted technologies in the writing process to enhance readability, improve language, and reduce word count within the specified limit. After using this tool/service, the authors reviewed and edited the content as needed and take full responsibility for the content of the publication.

### Consent for publication

All authors have read and approved the final version of the manuscript.

## Acknowledgements

We are grateful to the staff of the Consortium for Neglected Tropical Diseases and One Health at the Department of Biological Science, at UENR, Sunyani Ghana, for their support towards this research work. We thank the management of the Laboratory Department of the Berekum Holy Family Hospital for granting access and technical assistance. We also appreciate the efforts of the medical laboratory scientists in the microbiology department for their support throughout the benchwork phase of this study.

## Authors’ contributions

**J. K.A**., **G.D**., **R. D**., **N. A**., **& Y. B**.: Conception and design of study, Acquisition of data: laboratory or clinical, Analysis of data, Drafting of article and/or critical revision, Final approval of manuscript. **A.A**., **P-C. K**., **P. N**., **& D. O.K**.: Analysis of data, Drafting of article and/or critical revision, Final approval of manuscript. **K. B. O**.: Conception and design of study, Analysis of data, Drafting of article and/or critical revision, Final approval of manuscript.

## Availability of data and materials

The datasets used and/or analysed during this study are available from the corresponding author on reasonable request.

